# Emergence of SARS-CoV-2 Resistance with Monoclonal Antibody Therapy

**DOI:** 10.1101/2021.09.03.21263105

**Authors:** Manish C. Choudhary, Kara W. Chew, Rinki Deo, James P. Flynn, James Regan, Charles R. Crain, Carlee Moser, Michael Hughes, Justin Ritz, Ruy M. Ribeiro, Ruian Ke, Joan A. Dragavon, Arzhang C. Javan, Ajay Nirula, Paul Klekotka, Alexander L. Greninger, Courtney V. Fletcher, Eric S. Daar, David A. Wohl, Joseph J. Eron, Judith S. Currier, Urvi M. Parikh, Scott F. Sieg, Alan S. Perelson, Robert W. Coombs, Davey M. Smith, Jonathan Z. Li, for the ACTIV-2/A5401 Study Team

## Abstract

Resistance mutations to monoclonal antibody (mAb) therapy has been reported, but in the non-immunosuppressed population, it is unclear if *in vivo* emergence of SARS-CoV-2 resistance mutations alters either viral replication dynamics or therapeutic efficacy. In ACTIV-2/A5401, non-hospitalized participants with symptomatic SARS-CoV-2 infection were randomized to bamlanivimab (700mg or 7000mg) or placebo. Treatment-emergent resistance mutations were significantly more likely detected after bamlanivimab 700mg treatment than placebo (7% of 111 vs 0% of 112 participants, P=0.003). There were no treatment-emergent resistance mutations among the 48 participants who received bamlanivimab 7000mg. Participants with emerging mAb resistant virus had significantly higher pre-treatment nasopharyngeal and anterior nasal viral load. Intensive respiratory tract viral sampling revealed the dynamic nature of SARS-CoV-2 evolution, with evidence of rapid and sustained viral rebound after emergence of resistance mutations, and worsened symptom severity. Participants with emerging bamlanivimab resistance often accumulated additional polymorphisms found in current variants of concern/interest and associated with immune escape. These results highlight the potential for rapid emergence of resistance during mAb monotherapy treatment, resulting in prolonged high level respiratory tract viral loads and clinical worsening. Careful virologic assessment should be prioritized during the development and clinical implementation of antiviral treatments for COVID-19.

## INTRODUCTION

Across a broad spectrum of viral infections, host immune pressure^1,2^ and antiviral therapy^3-5^ can select for viral escape mutations. The detection and characterization of antiviral resistance mutations has been critical for the selection of appropriate antiviral therapies and to advance our understanding of viral adaptation against evolutionary pressures^6^. Monoclonal antibody (mAb) therapy is the current treatment of choice for non-hospitalized persons with early SARS-CoV-2 infections and mild to moderate COVID-19^7,8^. Bamlanivimab was the first mAb to receive FDA emergency use authorization (EUA) after it was demonstrated that treatment with bamlanivimab decreased nasopharyngeal (NP) SARS-CoV-2 detection and the risk of hospitalization or death when compared to placebo^9^. The emergence of SARS-CoV-2 sequence changes was reported shortly after the introduction of mAbs^7,10,11^, but there has not been definitive evidence that the emergence of SARS-CoV-2 resistance mutations can lead to altered *in vivo* intrahost viral replication dynamics and loss of therapeutic efficacy.

ACTIV-2/A5401 is a platform trial to evaluate efficacy of antiviral agents to prevent disease progression in non-hospitalized persons with symptomatic SARS-CoV-2 infection (NCT04518410). Participants were randomized to receive either bamlanivimab or placebo, with frequent NP swab and daily anterior nasal (AN) swab collection. We utilized quantitative viral load testing and Spike (S) gene next-generation sequencing to assess the emergence of viral resistance mutations to bamlanivimab and their impact on viral load dynamics. These results provide a window into the dynamic nature of SARS-CoV-2 intrahost viral population shifts and demonstrate for the first time that in non-immunosuppressed persons, the emergence of viral resistance can alter viral decay kinetics and lead to loss of antiviral activity.

## RESULTS

### SARS-CoV-2 resistance mutations emerging with mAb treatment were associated with changes in viral replication kinetics

A total of 94 participants were enrolled in the 7000mg cohort (48 in the treatment arm and 46 in the placebo arm, enrolled between August 2020 and October 2020) and 223 participants were enrolled in the ACTIV-2/A5401 phase 2 bamlanivimab 700mg cohort (111 in the treatment and 112 in placebo arms, enrolled between October 2020 and November 2020). The 7000mg dosing group was halted early due to the results of the BLAZE-1 study showing similar virologic efficacy between the bamlanivimab 7000mg and 700mg groups^9^. Viral sequences were successfully obtained from 207 participants in the 700mg bamlanivimab study and 78 in the 7000mg study from at least 1 respiratory sample with quantitative SARS-CoV-2 measurement ≥2 log_10_ RNA copies/mL at baseline or during 28 days of follow-up. Primary resistance mutations (L452R, E484K, E484Q, F490S and S494P)^7,12^ at ≥20% frequency were not detected in any participants in the 7000mg bamlanivimab study, either at baseline or following the single infusion. In the 700mg bamlanivimab arm, three participants had primary resistance mutations at baseline (one L452R in the setting of B.1.427/429/Epsilon variant infection and two participants with E484K) while the placebo arm had two cases of resistance mutations present at baseline (both L452R in the setting of Epsilon infection, Figure 1). Treatment-emergent mutations at ≥20% frequency (not detected at baseline) were significantly more likely to be detected after bamlanivimab 700mg treatment than placebo (7% vs 0%, P=0.003); E484K was found in 5 of 8 cases of emergent resistance, E484Q in two cases, and S494P in one case (Figure 1b). There were two cases of emerging resistance mutations present only as low frequency variants (<20% frequency): one participant in the placebo arm had an emergent F490S mutation (participant B2_9, Supplemental Figure 1), while one participant in the bamlanivimab 700mg treatment arm had emerging S494P concurrent with an emerging E484K (participant B2_6, Supplemental Figure 1).

**Figure 1:**
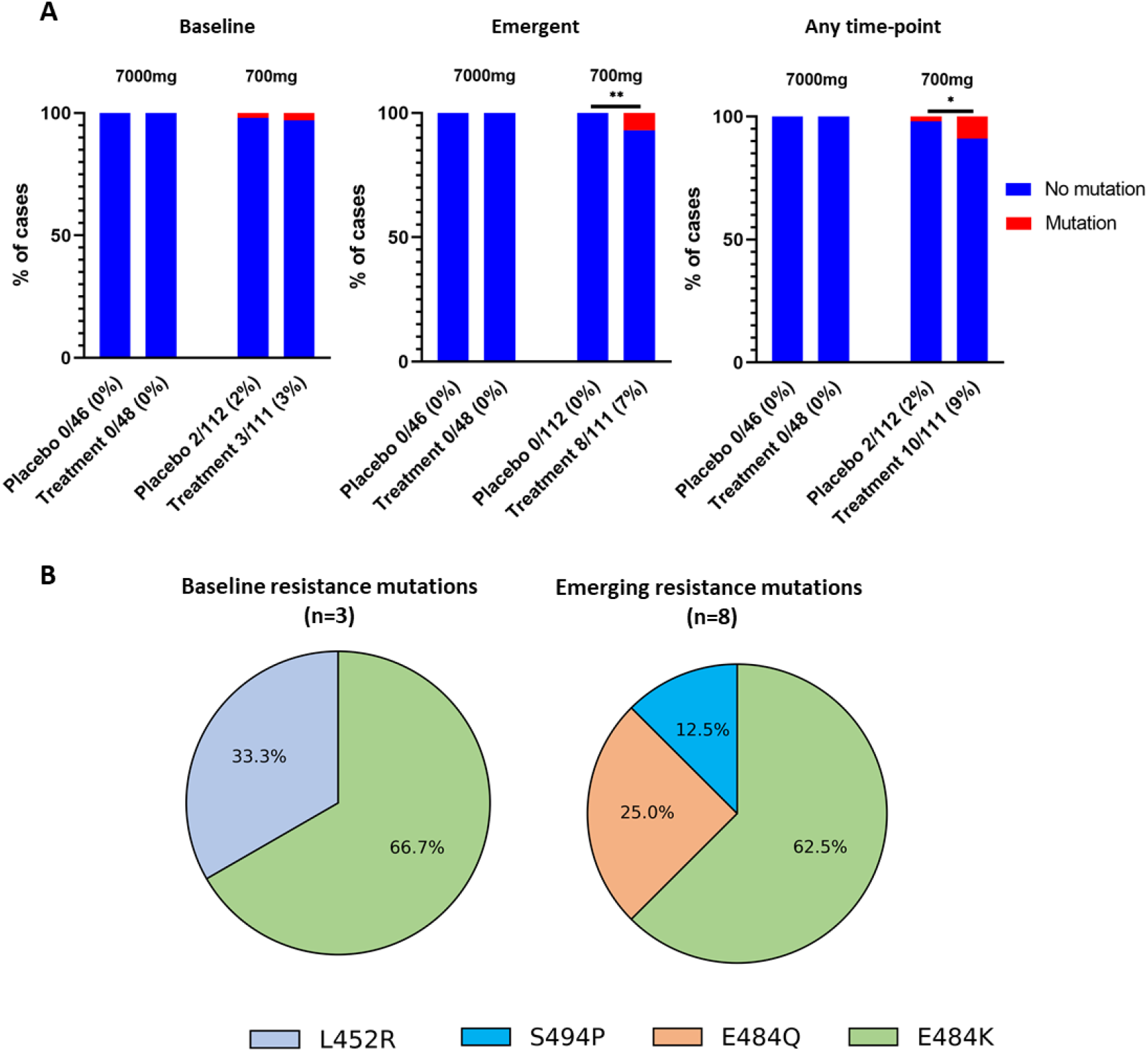
Prevalence of SARS-CoV-2 primary resistance mutations. (A) Percent of participants harboring primary resistance mutations L452R, E484K, E484Q, F490s and S494P at ≥20% frequency in the bamlanivimab 7000mg and 700 mg treatment and placebo arms at baseline, emergent and at any time-point. Participants without quantifiable viral load at baseline and/or follow-up time points were grouped with those without resistance. P-values were calculated using Fisher’s exact test. * P <0.05, ** P<0.01. (B) Pie-charts showing distribution of baseline and emergent resistance mutations in treatment arm. One participant had E484K at baseline with emerging E484Q mutation.

Quantitative SARS-CoV-2 viral loads were measured from NP swabs at days 0, 3, 7, 14, 21 and 28 of the trial, and from AN swabs daily at each of the first 14 days and at days 21 and 28. We assessed differences in viral loads in those receiving bamlanivimab 700mg treatment by the presence of emerging resistance. Pre-treatment NP and AN swab viral loads were higher for participants with emerging resistance mutations compared to those with no mutations (emerging vs no mutation viral loads at day 0, NP swab: median 7.6 vs 5.5 log_10_ copies/mL, P = 0.04; AN swab: median 6.6 vs 4.3 log_10_ copies/mL, P = 0.02, Table 1). Those with emerging resistance also had persistently elevated NP and AN viral loads throughout the first 14 days after study enrollment (Figure 2). Individuals with emerging resistance were older (emerging vs no mutation: median age 56 vs 45 years, P=0.01) and while not statistically significant, the median duration of symptoms at study entry was modestly shorter in those with emerging resistance compared to those without any mutations (emerging vs no mutations: median 4.5 vs 6.0 days, Table 1). Six of the participants with emerging resistance had samples available for baseline serologies and all were negative for IgG (Supplemental Figure 2). Measurements of bamlanivimab serum concentrations in participants with emerging resistance showed results generally concordant with expected values for the 700mg dose, including for maximal concentrations (Cmax) at the end of infusion and concentrations at day 28 (Supplemental Table 1)^13^. One participant (B2_6) did have a serum concentration at day 28 below the limit of quantitation and an elimination half-life faster than typical.

**Table 1:** Demographic characteristics of enrolled participants receiving bamlanivimab treatment comparing those with emerging resistance to those without any detected resistance mutations. One participant had both baseline and emerging resistance and was included in the Emerging resistance category. Statistical analysis was performed using Mann Whitney U tests for continuous variables and Fisher’s exact tests for discrete variables.

| **Characteristic** | **7000 mg Treatment w/**<br>**Bamlanivimab**<br>**(N = 48)** | **700 mg Treatment w/**<br>**Bamlanivimab**<br>**(No resistance)**<br>**(N = 101)** | **700 mg Treatment w/**<br>**Bamlanivimab**<br>**(Emerging**<br>**resistance)**<br>**(N = 8)** | **P-Value**<br>**(Comparison**<br>**between those**<br>**with and**<br>**without**<br>**resistance)** |
| --- | --- | --- | --- | --- |
| Age, median years [Q1,Q3] | 46 [ 33, 58] | 45 [34, 54] | 56 [50, 64] | 0.01 |
| Female sex, % | 54 | 50 | 50 | 1.0 |
| Race/Ethnicity, % |  |  |  |  |
| White | 56 | 85 | 75 | 0.61 |
| Black | 6 | 10 | 25 | 0.21 |
| Hispanic | 31 | 17 | 13 | 1.0 |
| Other | 6 | 5 | 0 | 1.0 |
| BMI, median score [Q1,Q3] | 28.2 [24.8, 31.8] | 28.2 [25.1, 33.7] | 29.4 [26.4, 38.2] | 0.36 |
| Baseline NP VL, median<br>log<sub>10</sub> SARS-CoV-2<br>copies/mL [Q1, Q3] | 5.2 [2.4, 6.6] | 5.5 [3.9, 6.8] | 7.6 [6.4, 8.0] | 0.04 |
| Baseline AN VL, median<br>log<sub>10</sub> SARS-CoV-2<br>copies/mL [Q1, Q3] | 4.1 [1.0, 5.9] | 4.3 [2.3, 6.1] | 6.6 [5.8, 7.3] | 0.02 |
| Days from symptom onset to<br>randomization, median days<br>[Q1, Q3] | 6.0 [4.0, 8.0] | 6.0 [5.0, 8.0] | 4.5 [2.5, 7.5] | 0.15 |

**Figure 2:**
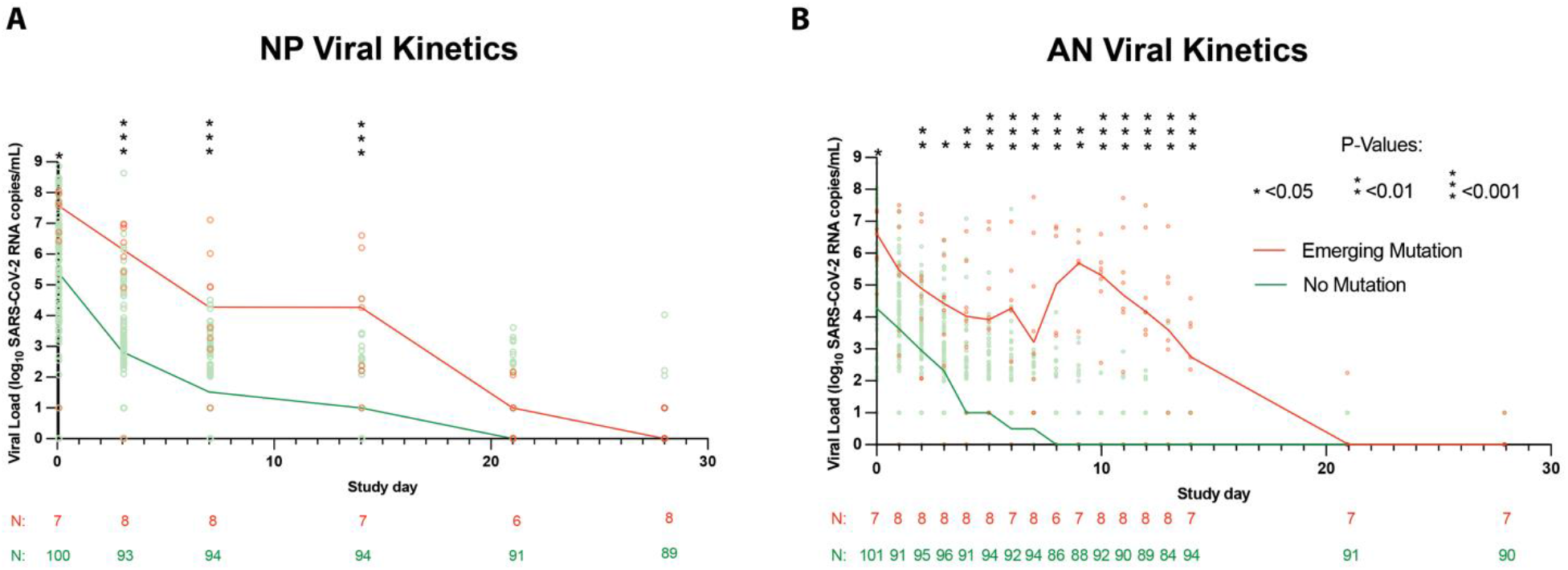
SARS-CoV-2 viral kinetics in the bamlanivimab 700mg treatment arm. SARS-CoV-2 viral loads from (A) Nasopharyngeal (NP) swabs (collected at day 0, 3, 7, 14, 21 and day 28) and (B) from anterior nasal (AN) swabs (collected daily through day 14 followed by day 21 and day 28) plotted against study day. Lines show median viral load. Viral loads between groups were compared at each time point using the Mann-Whitney U tests denoted by asterisks wherever significant. The lower limit of quantification was 2.0 log_10_ SARS-CoV-2 RNA copies/mL while the lower limit of detection was 1.0 log_10_ copies/mL.

### Evidence of dynamic SARS-CoV-2 viral population shifts and differential viral fitness after mAb treatment

We observed that the emergence of SARS-CoV-2 resistance mutations was closely associated with a relatively consistent change in viral load kinetics. This is exemplified in Figure 3 with two examples of viral rebound in participants with emergence of escape variants. In case B2_3, intensive S gene sequencing of virus isolated from the AN swabs revealed the emergence of the E484K resistance mutation on study day 3 as a low frequency variant that rapidly took over as the majority population by the next day (Figure 3A, lower panel) and was associated with a 3.6 log_10_ increase in AN swab viral loads over the subsequent 4 days to a peak of 7.8 log_10_ copies/mL on study day 7 before declining. For B2_2, the participant had evidence of baseline E484K mutation and low-frequency E484Q in the NP swab (Figure 3B). The AN swab showed low frequency E484K and Q mutations. After bamlanivimab treatment, there were rapid, dynamic shifts in the viral population in the AN swab sample including both the E484K and Q mutations, with the viral load peaking at 6.8 log_10_ copies/mL over 8 days concurrent with E484Q becoming the dominant mutation. Among the 8 participants with emerging resistance, the median AN swab viral load increase was 3.3 (range 0.3 - 5.2) log_10_ RNA copies/mL over a median of 4.5 days (Supplemental Figure 1) and this viral rebound is highlighted in the comparison of median viral loads between those with and without emerging resistance mutations (Figure 2).

**Figure 3:**
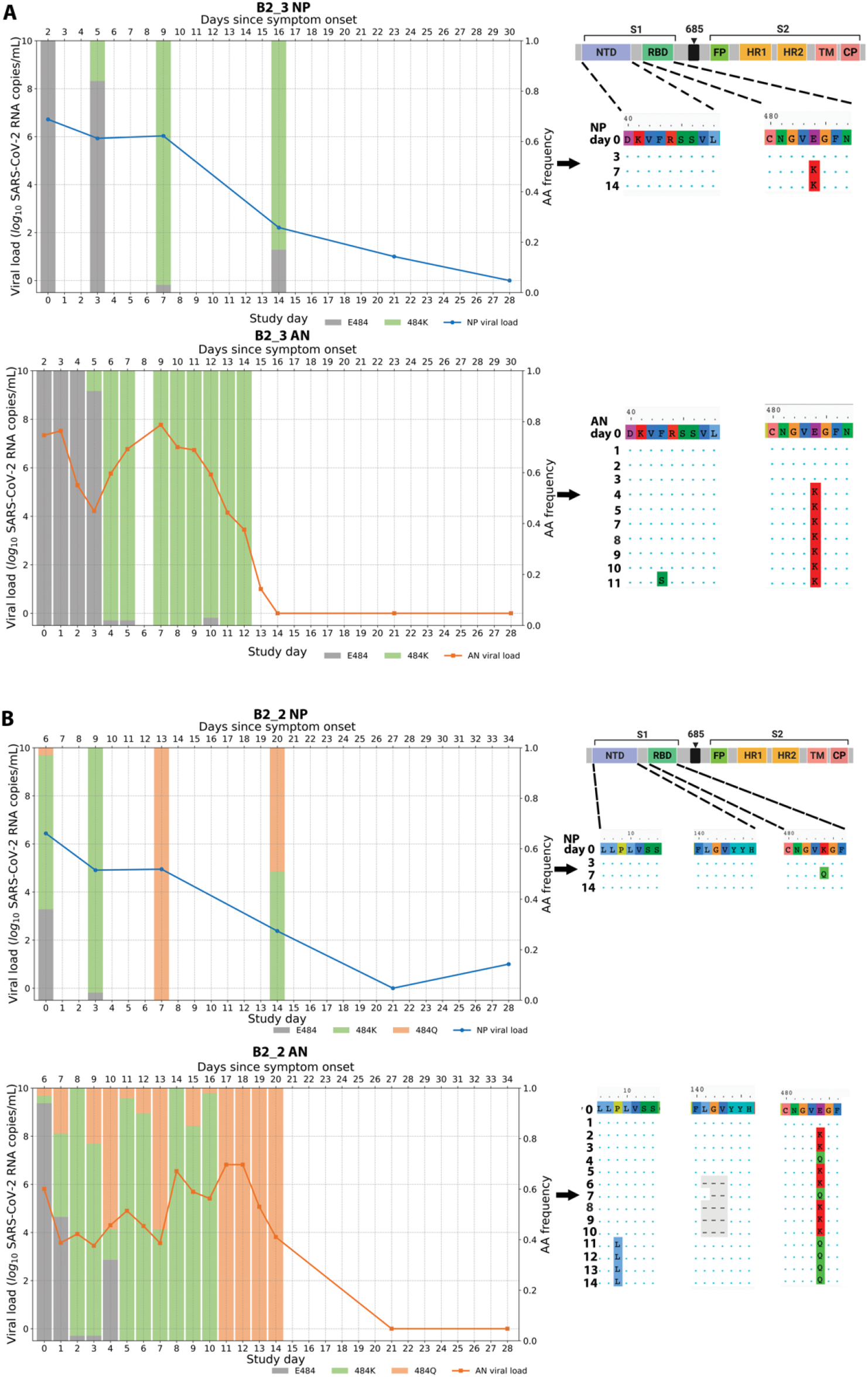
Evidence of viral rebound and/or slow viral decay coupled with dynamic viral population shift and potential compartmentalization. Viral RNA from nasopharyngeal (NP) swabs and anterior nasal (AN) swabs were sequenced and results from two example participants from the bamlanivimab 700mg treated group are shown. (A) Participant B2_3 showed emergence of E484K and viral rebound between study days 3 and 7. (B) Participant B2_2 showed emergence of a mixed population of E484K and E484Q viruses along with multiple rebounds and slow viral decay. Alignments of consensus sequences from both compartments show position of primary escape and other consensus-level mutations at each time point. CP denotes cytoplasmic domain, FP fusion peptide, HR1 heptad repeat 1, HR2 heptad repeat 2, NTD N-terminal domain, RBD receptor binding domain, S1 subunit 1, S2 subunit 2, and TM transmembrane domain.

To quantify the replicative fitness of the different strains, we developed a mathematical model and fit it to both viral load data and variant frequency data collected from 6 participants in the treatment arm with either E484K or Q resistance emergence. In this model, we assumed that each variant is initially present and grows or declines exponentially at a constant rate (see Methods) as was consistent with the data (Supplemental Figure 3). We chose the first 8-13 time points for model fitting covering the emergence of the resistance mutations but prior to the eventual viral load declines. We estimated both the initial viral load and the rate of exponential increase or decrease for each variant. From the estimation across the 6 individuals, the wild-type amino acid (i.e., 484E), always declined under treatment, with the exponential rate varying from -0.2 to -3.2 per day (Supplemental Table 2). In contrast, the mutant 484K always increased under mAb treatment, confirming it was a resistant mutant, with an exponential growth rate that varied over a wide range (0.5 to 2.3 per day, Supplemental Figure 3G). The 484Q mutant was found in two participants, including low-frequency E484K and Q present at baseline in the AN swab sample of participant B2_2. We estimated that virus harboring 484Q was more fit than 484K in the setting of antibody treatment and grew at approximately twice the rate of the 484K variant (Supplemental Table 2). In participant B2_5, viral loads in the setting of the 484Q mutant declined, but with a rate much slower than the wildtype 484E, suggesting that it is more fit than 484E in the presence of the mAb or developing host immune responses.

### Emergence of additional Spike polymorphisms

In those with emerging bamlanivimab resistance, we next assessed the emergence of additional S gene sequence changes outside of the primary sites of resistance (L452R, E484K, E484Q, F490S and S494P). We found that emergence of additional polymorphisms was common and could be detected in all participants with either baseline or emerging bamlanivimab primary resistance mutations, although most were present at low-frequencies (Figure 4). One emergent polymorphism, Q493R, was detected in B2_7 and has been described as a potential bamlanivimab resistance site^10^. Interestingly, a number of polymorphisms were detected at sites distinct from the bamlanivimab site of activity and likely reflect escape from host immune pressures. For example, deletions at amino acid positions 141-143 were detected to emerge in both participants B2_2 and B2_4. These N-terminal domain (NTD) deletions have previously been described in immunosuppressed participants with immune escape and persistent COVID-19^14,15^. These deletions have also been detected in the wider pandemic and represent a common site of viral escape against antibody pressure on the NTD^16^. A number of emerging polymorphisms were also detected that are also in several variants of concern and postulated to be involved in either immune escape or enhanced receptor binding. These include L5F (PID B2_2, also in B.1.526/Iota), P9L (PID B2_2, also in C.1.2), L18F (PID B2_8, also in B.1.351/Beta, P.1/Gamma), D138Y (in P.1/Gamma), N501Y (ACE-2 binding domain mutation in PID B2_10, also in B.1.1.7/Alpha, B.1.351/Beta, P.1/Gamma), and P681H (furin cleavage site mutation in PID B2_10, also in B.1.1.7/Alpha)^17,18^.

**Figure 4:**
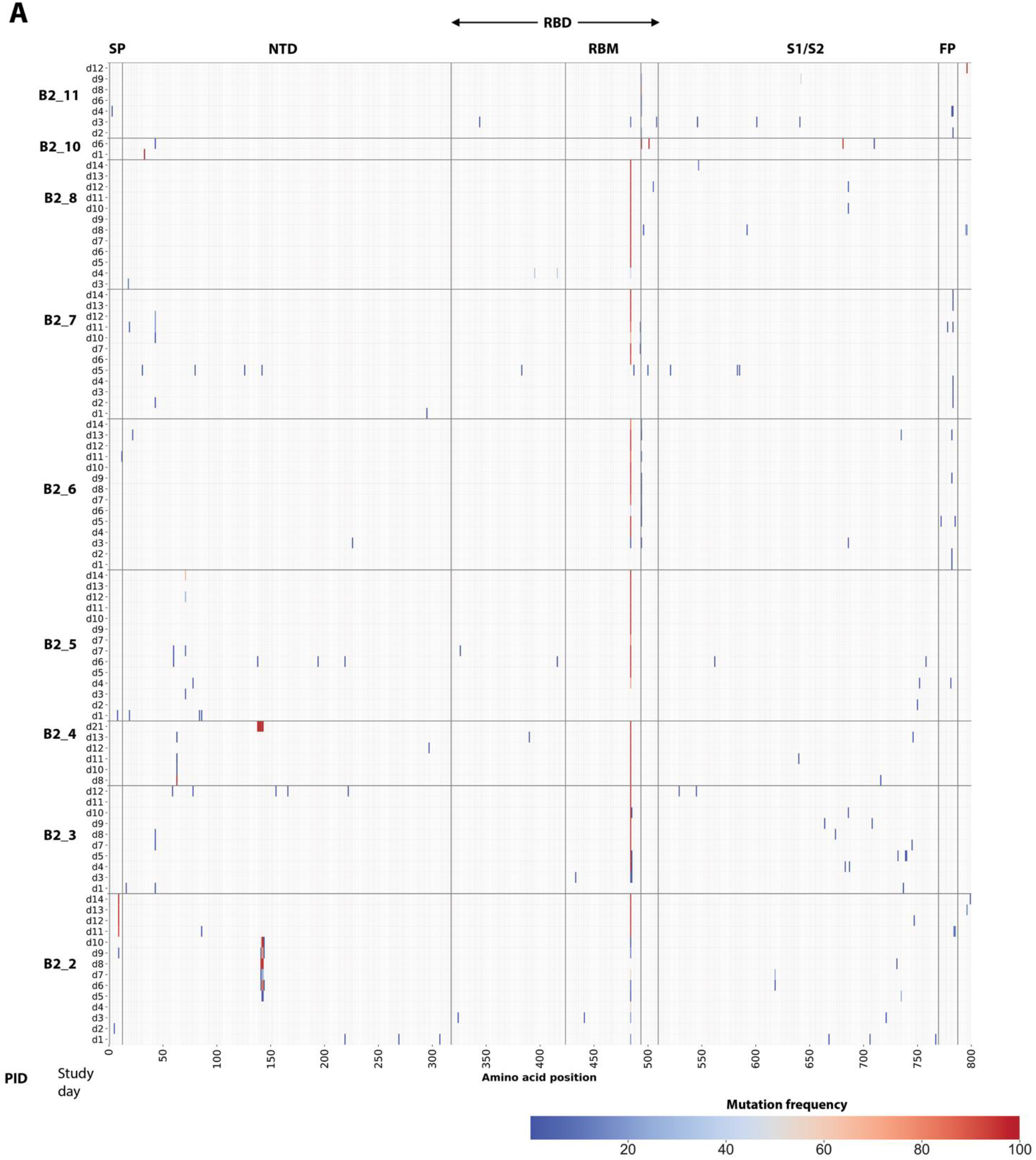
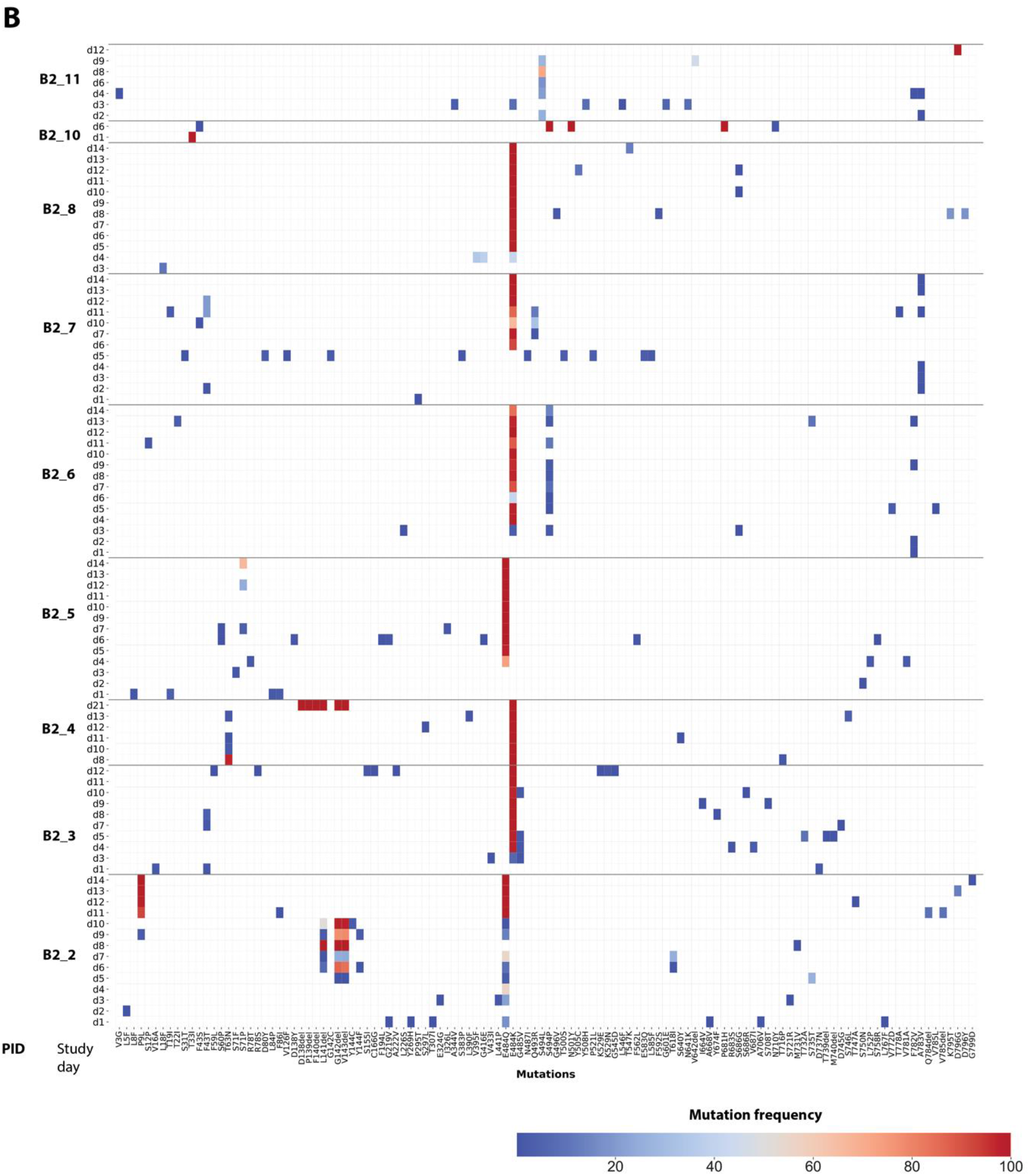
Heat map showing distribution of Spike polymorphisms from AN swab samples in bamlanivimab 700mg treated participants with emerging primary resistance mutations. (A) Panel A shows polymorphism in context of near-full length Spike gene. Y-axis shows participants’ ID followed by day of sample collection, while x-axis shows amino-acid positions in Spike gene. Different domains of Spike are shown at the top while color indicates frequency of polymorphisms starting with blue indicating lowest value while red indicates highest value in the scale. (B) Zoomed-in heat-map showing sites which harbors polymorphisms at least one of the samples across different participants. The order of samples is same that in panel A while x-axis denotes amino-acid sites with number indicating position of amino-acids while letter before and after the numbers indicate wild-type and polymorphic amino-acid respectively. SP denotes signal peptide, NTD N-terminal domain, RBD receptor binding domain, RBM Receptor binding domain, S1 subunit 1, S2 subunit 2, and FP fusion peptide.

We used day 7 NP swab sequencing results to compare the rate of polymorphism emergence across the participant groups as AN swab sequencing was performed only for participants with evidence of resistance emergence. We detected no significant differences in the number of emerging polymorphisms between those with emerging resistance, treated participants without resistance and participants who received placebo (Supplemental Figure 4).

### Viral rebound during after bamlanivimab resistance emergence is associated with worsened symptoms

To assess the clinical impact of the viral load resurgence seen in those with emerging bamlanivimab resistance mutations, we compared longitudinal symptom scores for bamlanivimab-treated participants with and without emerging resistance mutations. Total symptom scores were calculated based on a 28 day diary completed by the participants for 13 targeted symptoms^19^. On an individual-level, higher symptom scores were frequently detected after the emergence of resistance and increase in respiratory tract viral loads (Figure 5A). In the population analysis, there was no significant differences in symptom scores between groups before the emergence of resistance mutations. In participants of the bamlanivimab 700mg treatment arm with emerging resistance mutations, median AN viral load increase began at the time of resistance detection, with significantly higher subsequent viral loads and total symptom scores compared to those in the treatment arm without resistance (Figure 5B).

**Figure 5.**
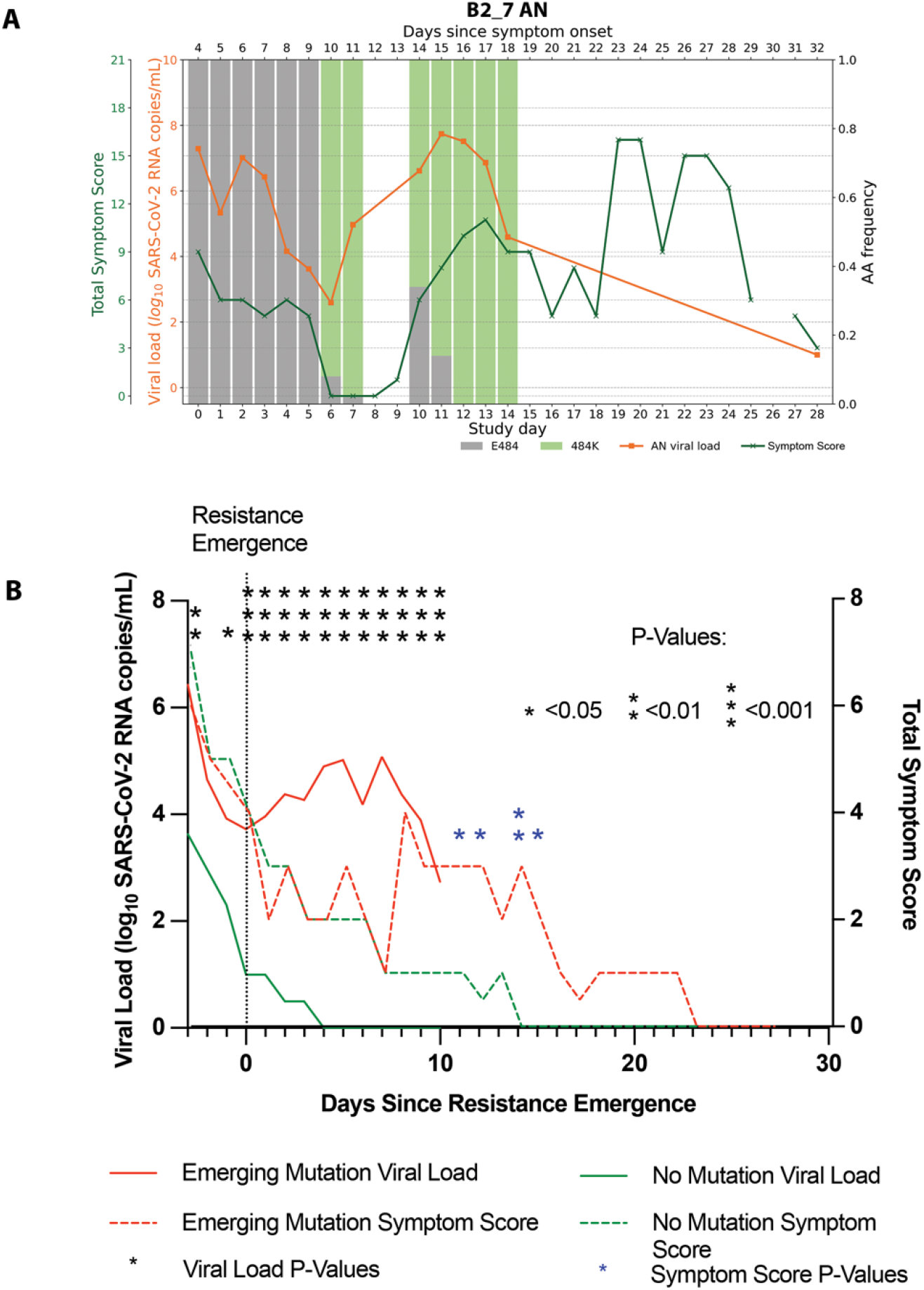
Worsened COVID-19 symptoms with viral resurgence after emergence of resistance mutations. (A) Example of increased anterior nasal (AN) viral load and total symptom score trend for one participant, B2_7, with emerging E484K resistance after bamlanivimab 700mg treatment. (B) Median AN viral load (solid line) and total symptom score (dashed line) plotted from the time of resistance emergence to ≥20% of the viral population (day 0) for participants in the bamlanivimab 700mg treatment group with (red) and without (green) emerging resistance mutations. For participants without emerging resistance, day 0 was equivalent to study day 4, which represented the median day of resistance emergence for those with emerging resistance. Symptom score between the emerging resistance and no emerging resistance groups is compared at each day by Mann Whitney U tests. * P < 0.05, ** P < 0.01.

## DISCUSSION

In this virologic analysis of a randomized placebo-controlled clinical trial of non-hospitalized persons with early COVID-19, we report the emergence of resistance mutations to the mAb bamlanivimab and the effects of these mutations on viral decay and clinical symptoms. These results represent the clearest evidence to date of several key principles: 1) the dynamic nature of SARS-CoV-2 evolution and replication during mAb treatment, 2) treatment-emergent SARS-CoV-2 resistance mutations alter viral replication kinetics and extend the period of high viral loads, 3) drug resistance mutations can adversely affect both the virologic and clinical efficacy of a COVID-19 antiviral medication, and 4) the emergence of resistance with mAb treatment is dependent on the treatment dose.

In immunocompromised persons with COVID-19, viral evolution can lead to immune escape and rapid emergence against even combination mAb therapy^11,15,20,21^. Whether these findings are generalizable to the immunocompetent population has been unclear and there has not been definitive evidence that the emergence of SARS-CoV-2 escape mutations impacts *in vivo* viral replication dynamics and loss of therapeutic efficacy. In this study of bamlanivimab in a general population of outpatients with mild to moderate COVID-19, we show that resistance mutations to monoclonal antibody treatment can emerge quickly and are associated with rapid and sustained increase in respiratory tract SARS-CoV-2 viral load. Importantly, this increase in viral load was followed by significantly worsened clinical symptoms over the subsequent days. These results are consistent with previous reports that during acute SARS-CoV-2 infection, high-level respiratory tract viral loads often precede symptom onset by 1-2 days^22^.

We were able to identify several potential factors that may increase or decrease the risk of mAb resistance. We found that older age and higher baseline respiratory tract viral load were associated with higher risk of resistance emergence, while none of the 48 participants treated with the higher dose bamlanivimab 7000mg therapy developed resistance. Studies of mAb treatments have suggested that earlier initiation of therapy during periods of high respiratory tract viral load is associated with a greater virologic response and likely improved therapeutic efficacy^23^. Our data suggest that mAb treatment during periods of high-level viral loads may come at the cost of increased risk of resistance emergence, although this effect may be mitigated by using higher doses of mAbs. Interestingly, we also noted frequent increase in viral loads associated with resistance emergence that lasted several days to more than a week before declining. Such prolonged rise in viral loads is unusual, especially as these individuals had a median of 5 days of symptoms by the time of study entry and it’s expected that levels of respiratory viral loads should already be declining^24^. While the exact cause is unclear, this finding raises several intriguing possibilities. First, antiviral mAb therapies may have host immune modulating effects beyond their capacity to bind and neutralize viral particles^25^. It is unknown whether mAb therapeutics may in some cases interfere with host immune responses, especially in the setting of mAb resistance, leading to suboptimal viral control. Alternatively, there have also been reports from *in vitro* studies that certain SARS-CoV-2-specific antibodies may lead to antibody-dependent enhancement of infection through an Fcγ receptor-dependent mechanism^26^, particularly at sub-neutralizing concentrations, although *in vivo* confirmation has been challenging to obtain.

Our study also found that SARS-CoV-2 populations can turn over quickly, allowing for quick selection of drug resistance associated mutations. In fact, viral populations were found to be able to completely shift from fully sensitive to fully resistant viruses within 24 hours. The emerging primary resistance mutations (e.g., E484K/Q) described in this report not only confer resistance to mAb therapy but can also lead to decreased efficacy of vaccine-induced immune responses^27^. While the rate of total polymorphism accumulation did not appear to be higher in those who developed bamlanivimab resistance, many of the emerging polymorphisms are also key mutations found in several variants of concern/interest (VOC/VOIs, including B.1.1.7/Alpha, B.1.351/Beta and P.1/Gamma), which are associated with increased transmission efficiency and enhanced outbreaks^28^. The impact of treatment-induced resistance mutations on the spread of these key escape mutations should be further assessed.

One limitation to this study is that bamlanivimab is no longer used clinically as a single agent. These results, though, provide important proof of principle for the role that drug resistance may have on virologic and clinical efficacy of SARS-CoV-2 antiviral therapies. These lessons have implications for the development of novel COVID-19 antiviral therapies and have continued relevance for other clinically-approved single agent monoclonal antibody treatments^29^ and for combination therapies where one agent may be ineffective due to circulating variants^13^. Another limitation of this study is the limited sample size of this phase 2 study, especially in the bamlanivimab 7000mg cohort. While treatment-emergent mutations were not found in ACTIV-2 participants receiving the higher 7000mg dose of bamlanivimab, they were frequently detected in the larger BLAZE-1 phase 2 trial of the 7000mg dose^7^. One difference between these studies was the longer duration since symptom onset for the ACTIV-2 participants, who enrolled a median of 6 days since symptom onset versus 4 days for the BLAZE-1 participants. This likely led to higher pretreatment viral loads, which we found to be a risk factor for resistance emergence. Unfortunately, baseline viral loads could not be compared between studies as the BLAZE-1 study did not use a quantitative SARS-CoV-2 viral load assay. These disparate results highlight the importance of incorporating quantitative viral load testing and resistance testing for COVID-19 treatment trials of mAbs and other antiviral agents.

In summary, these results provide clear evidence that mAb treatment can rapidly select for SARS-CoV-2 resistance, leading to dramatic viral rebound and worsened symptom severity. While initiation of mAb treatment during early infection is recommended for optimal therapeutic benefit, our results suggest that emerging resistance is a potential risk with treatment during periods of high level viral replication. These findings have implications for the design and utilization of SARS-CoV-2 antiviral therapeutics and provide insights into the prevention of SARS-CoV-2 resistance. Careful virologic and pharmacologic assessment of new treatments for COVID-19 should be prioritized.

## METHODS

### Study participants and sample collection

The study participants were enrolled in the ACTIV-2/AIDS Clinical Trials Group (ACTG) A5401 phase 2 randomized, placebo-controlled trial of bamlanivimab 7000mg and 700mg mAb therapy. Symptomatic adults ≥18 years of age with a documented positive SARS-CoV-2 antigen or nucleic acid test were enrolled if the diagnostic sample was collected ≤7 days prior to study entry and within 10 days of symptom onset. The 7000mg dosing group was halted early due to the results of the BLAZE-1 study showing similar virologic efficacy between the bamlanivimab 7000mg and 700mg groups^9^. A total of 95 participants were randomized in the bamlanivimab 7000mg study and 222 participants were randomized in the 700mg study and received an intervention (one bamlanivimab or placebo intravenous infusion). All participants provided written informed consent. Nasopharyngeal (NP) swab samples were collected by research staff at study days 0, 3, 7, 14 and 28, while anterior nasal (AN) swabs were self-collected by participants daily through study day 14 and at days 21 and 28. Swabs were placed in 3ml of media (RPMI with 2% FBS).

Total symptom scores were calculated based on a 28 day diary completed by the participants for 13 targeted symptoms^19^. The targeted symptoms are feeling feverish, cough, shortness of breath or difficulty breathing, sore throat, body pain or muscle pain or aches, fatigue, headache, chills, nasal obstruction or congestion, nasal discharge, nausea, vomiting, and diarrhea. Each symptom is scored daily by the participant as absent (score 0), mild (1) moderate (2) and severe (3).

### SARS-CoV-2 viral load testing and S gene next-generation sequencing

SARS-CoV-2 viral load from NP and AN swab samples were quantified using the Abbott m2000 system. SARS-CoV-2 quantitative Laboratory Developed Test (LDT) was developed utilizing open mode functionality on *m*2000*sp/rt* (Abbott, Chicago, IL) by using EUA Abbott SARS-CoV-2 qualitative reagents^30^. Identical extraction and amplification protocols developed for RealTi*m*e SARS-CoV-2 qualitative EUA assay were also used for the development of the RealTi*m*e SARS-CoV-2 quantitative LDT^31^. In this assay, 2 calibrator levels (3 log_10_ RNA copies/mL and 6 log_10_ RNA copies/mL) tested in triplicate were used to generate a calibration curve and 3 control levels (negative, low positive at 3 log_10_ RNA copies/mL and high positive at 5 log_10_ RNA copies/mL) were included in each run for quality management. In addition, batches of a matrix-specific control (“external” swab control) with a target of 200 copies per mL were prepared and one unit was included in every run. All controls were monitored using Levy-Jennings plots to monitor inter-run precision. Specimens that were greater than 7 log_10_ RNA copies/mL were diluted 1:1000 and rerun to obtain an accurate viral load result. The lower limit of quantification was 2.0 log_10_ SARS-CoV-2 RNA copies/mL.

S gene sequencing was performed on NP swab samples at two time points for all participants: baseline (study entry) and the last sample with a viral load (VL) ≥ 2 log_10_ SARS-CoV-2 RNA copies/mL. In participants with evidence of slow viral decay (VL ≥ 2 log_10_ copies/mL at study day 14) or viral rebound (increase in NP swab VL), we performed S gene sequencing of all NP swab samples with VL ≥ 2 log_10_ copies/mL. Sequencing of daily AN swab samples was performed for participants with any emerging bamlanivimab resistance mutations detected on NP swab samples. Viral RNA extraction was performed on 1 mL of swab fluid by use of the TRIzol-LS™ Reagent (ThermoFisher), as previously described^32^. cDNA synthesis was performed using Superscript IV reverse transcriptase (Invitrogen), per the manufacturer’s instructions. Spike gene amplification was performed using a nested PCR strategy with *in-house* designed primer sets targeting codons 1-814 of Spike. PCR products were pooled, and Illumina library construction was performed using the Nextera XT Library Prep Kit (Illumina). Sequencing was performed on the Illumina MiSeq platform. Raw sequence data were analyzed using PASeq v1.4 (https://www.paseq.org). Briefly, data were quality filtered using Trimmomatic (v0.30), using a Q25/5 bp sliding window and a 70 bp minimum length. Non-viral contamination was filtered out using BBsplit v35.76. Filtered reads were then merged with pear v0.9.6 aligned to the reference sequence using Bowtie2 v2.1.0). Amino acid variants were then called at the codon level using perl code and used for resistance interpretation with a 1% limit of detection.

### Detection of Spike mutations

We assessed the presence of previously confirmed bamlanivimab resistance mutations (L452R, E484K, E484Q, F490S, and S494P)^7,12^. The detection of resistance mutations down to 1% frequency was performed using Paseq^33^. Mutations detected by next-generation sequencing at <20% of the viral population were labelled as “low frequency” variants as they would largely be missed by traditional Sanger sequencing. A minimum average of 500x sequencing coverage per sample was required for variant calling.

### Serology

IgG antibodies recognizing SARS-CoV-2 Spike (S), Receptor binding domain (RBD), Nucleocapsid (N) and N terminal domain (NTD) proteins were measured in serum samples using a commercially available multiplex kit (catalog # K15359U) from Meso Scale Diagnostics (MSD, Rockville, MD). Assays were performed according kit instructions. Briefly, plates were treated with MSD Blocker A to prevent non-specific antibody binding. Serum samples were thawed and diluted 1:500 and 1:5000 in MSD diluent. IgG was detected by incubation with MSD SULFO-TAG anti-IgG antibody. Measurements were performed with a MESO Quickplex SQ 120 reader. Three internal serum controls provided by MSD were run with each plate. Pre-pandemic sera from healthy adult donors (n=10; AMSBIO LLC, Cambridge MA 02141) were included as additional negative controls for the assays. Threshold values for antibody titers (S, RBD and N proteins) were provided by MSD and were based on analyses of 200 pre-2019 and 214 COVID+ (PCR-confirmed) COVID-9 patients. The thresholds utilized provide 84%, 71% and 71% sensitivity and 99.5%, 98.5% and 100% specificity for Spike, RBD and N antibody responses, respectively. NTD thresholds were not available.

### Pharmacokinetic analysis

Blood samples for quantitation of bamlanivimab serum concentrations were collected pre-dose and at the following times after the end of infusion: 30 minutes, days 14 and 28 and weeks 12 and 24. Pharmacokinetic parameters of interest were maximum concentration (Cmax), elimination half-life and clearance (CL) and were calculated based on the statistical moment theory using the trapezoidal rule and linear regression (WinNonLin, Certara, Princeton, NJ, USA).

### Statistical analysis and mathematical modeling

The non-parametric Mann-Whitney U test was used to compare differences in viral loads between groups. Chi-squared tests and Fisher’s exact tests were used for analyses of proportions. All statistical analyses were performed in GraphPad Prism (Version 9.1.1). Intensive AN swab viral loads and sequences were used for mathematical modeling. The mutational load was calculated by multiplying resistance mutation frequency by the total viral load. In the model, we assumed that the *i*th variant, *V*_i_, has an initial load *V*_i,0_ and its population size changes exponentially at a constant rate, *r*_i_:

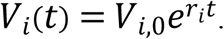

Then, the total viral load at time t, *V*(*t*) was calculated as:

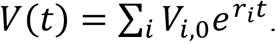

The model predicted frequency of each variant was

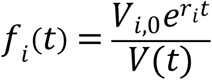

We estimated the initial load, *V*_i,0_ and the rate of exponential increase/decrease, *r*_i_, from the viral load and viral frequency data fitted simultaneously. Note that in this model, we assumed that the observed mutants were present at the time of antibody infusion or were produced quickly near that time.

To calculate the goodness of fit of the model to the data, we first calculated the residual sum of squares (RSS) between the model predicted viral load and the measured viral load on a logarithmic scale using log10. The log-scale was used because viral loads were measured using PCR and thus the measurement error was multiplicative, making the logarithm the natural scale to use. We then calculated the RSS between model predicted frequencies and measured frequencies for the mutants. The final RSS was calculated as the sum of the two RSS errors:

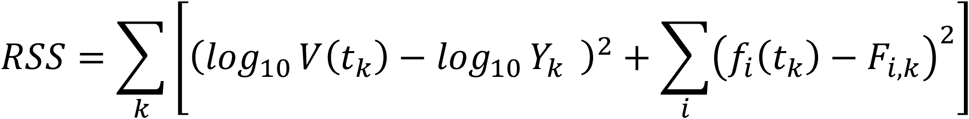

Where *k* denotes the *k*^th^ time point, *Y*_k_ and *F*_i,k_ denote the measured viral load and the measured frequency for the *i*^th^ variant at the *k*^th^ time point. All data will be available by request to the AIDS Clinical Trials Group and code will be available upon request to the authors.

## Data Availability

Data available subjected to author's consent.

## Author contributions

MCC, KC, RC, DMS, JZL conceptualized and performed the study, MCC, RD, JPF, JR, CC performed resistance analysis experiment; RMR, RK, ASP performed mathematical modeling; JD, AG performed viral load analysis; UP, SS performed serological analysis; CM, MH, JR performed statistical analysis.

## Acknowledgements

We would like to thank the participants, site staff, site investigators, and the entire ACTIV-2/A5401 study team. We thank the PASeq team (Drs. Roger Paredes and Marc Noguera Julian) for their support.

## Notes

**Financial support:** This study was supported by the National Institute of Allergy and Infectious Diseases; ACTIV- 2/A5401 ClinicalTrials.gov number NCT04518410. Portions of this work were performed under the auspices of the U.S. Department of Energy through Los Alamos National Laboratory (LANL), which is operated by Triad National Security, LLC for the National Nuclear Security Administration of the U.S. Department of Energy (contract No. 89233218CNA000001). The work was also supported by LANL’s Laboratory Directed Research and Development program (projects No. 20200743ER, 20200695ER, and 20210730ER), by NIH grants R01-AI028433, R01-OD011095 (ASP), R01-AI15270301 (RK) and R01-AI116868 (RMR), by the Defense Advanced Research Projects Agency (contract No. HR0011938513) and by the DOE Office of Science through the National Virtual Biotechnology Laboratory, a consortium of DOE National Laboratories focused on response to COVID-19, with funding provided by the Coronavirus CARES Act.

**Disclosures:** KWC has received research funding to the institution from Merck Sharpe & Dohme. PK and AN are employees and shareholders of Eli Lilly. ALG reports contract testing from Abbott and research support from Merck and Gilead. ESD has consulted for Gilead, Merck and ViiV and received research support from Gilead and ViiV. ASP has consulted for Amphylx Pharmaceticals. DMS has consulted for Bayer Pharmaceuticals, Linear Therapies, Matrix Biomed, FluxErgy and Brio Clinical. JZL has consulted for Abbvie.

### Competing Interest Statement

KWC has received research funding to the institution from Merck Sharpe & Dohme. PK and AN are employees and shareholders of Eli Lilly. ALG reports contract testing from Abbott and research support from Merck and Gilead. ESD has consulted for Gilead, Merck and ViiV and received research support from Gilead and ViiV. ASP has consulted for Amphylx Pharmaceticals. DMS has consulted for Bayer Pharmaceuticals, Linear Therapies, Matrix Biomed, FluxErgy and Brio Clinical. JZL has consulted for Abbvie.

### Clinical Trial

NCT04518410

### Funding Statement

This study was supported by the National Institute of Allergy and Infectious Diseases ACTIV-2/A5401 ClinicalTrials.gov number NCT04518410.

### Author Declarations

Ethics oversight was performed by a central IRB (Advarra). Informed consent was obtained from all participants.

