## Supplemental file for "Emergence of SARS-CoV-2 Resistance with Monoclonal Antibody Therapy"

**Supplemental Figure 1**. Viral loads and frequencies of primary resistance mutations from nasopharyngeal swab (NP) and anterior nasal swab (AN) samples for participants displaying primary bamlanivimab resistance mutations.

### Placebo arm participants with baseline resistance

| Participant ID | Nasopharyngeal | Anterior Nasal |
| --- | --- | --- |
| B2_12 | | |
| B2_13 | | |

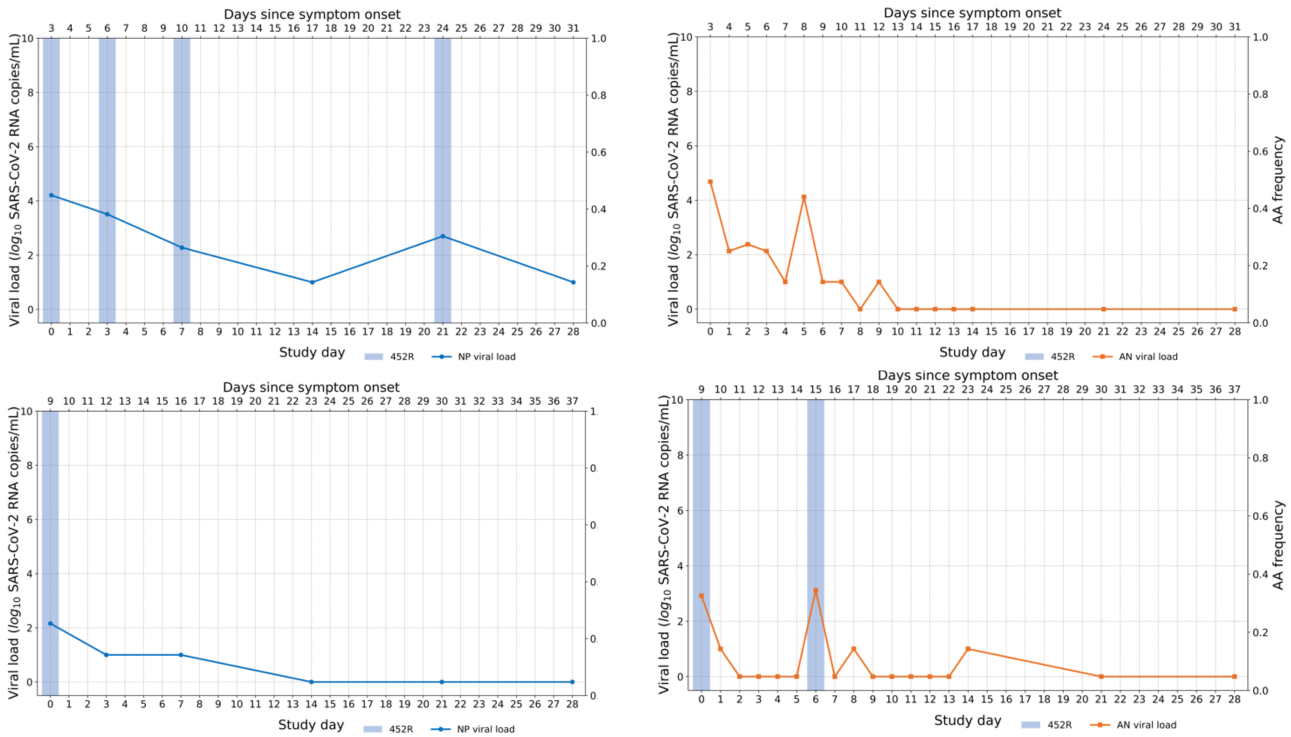


### Placebo arm participant with emerging resistance

| Participant ID | Nasopharyngeal | Anterior Nasal |
| --- | --- | --- |
| B2_9 | | |

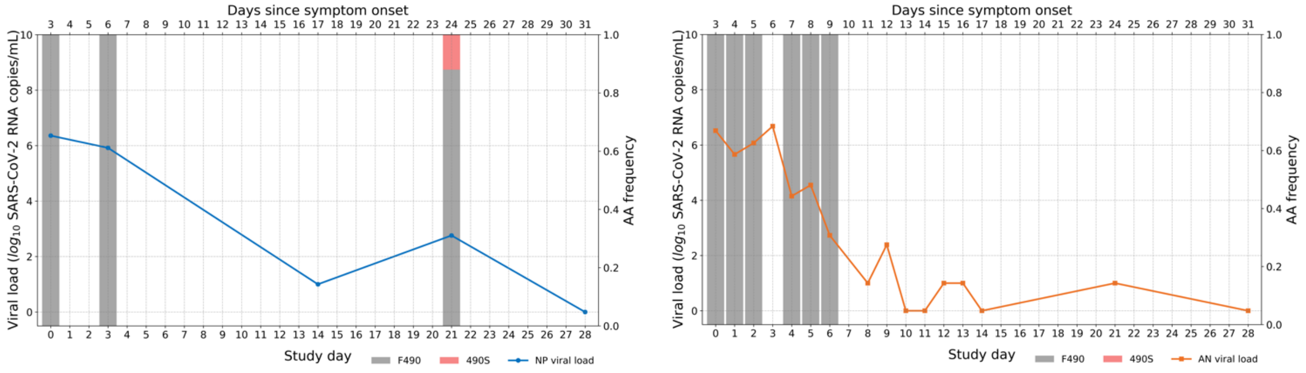

**Treatment arm participants with baseline resistance**

| Participant ID | Nasopharyngeal | Anterior Nasal |
|---|---|---|

B2_1

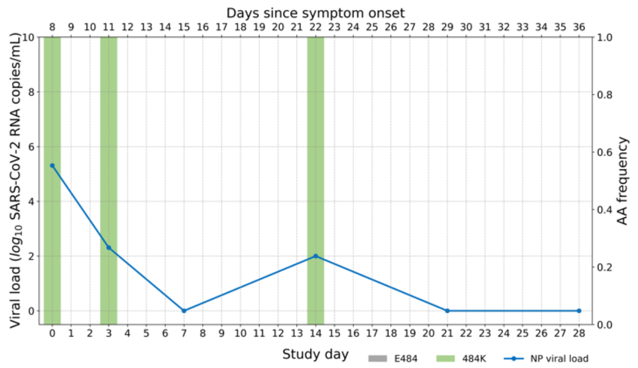


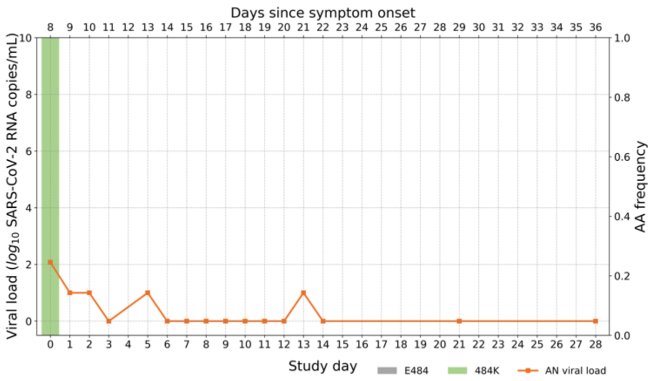


B2_11

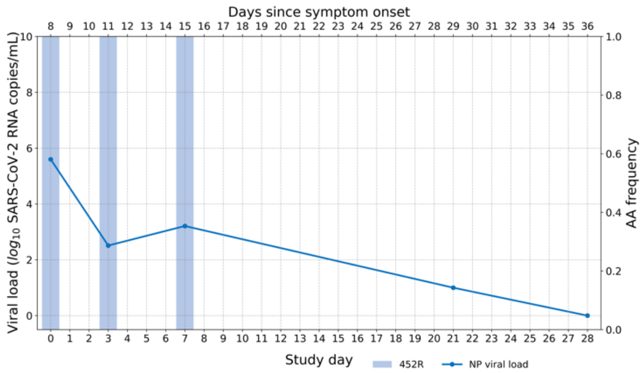


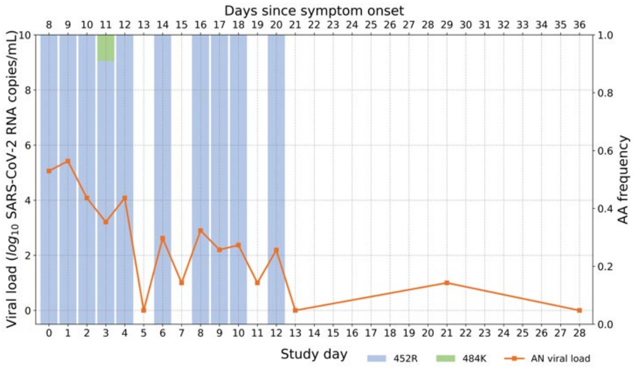


**Treatment arm participants with emerging resistance**

| Participant ID | Nasopharyngeal | Anterior Nasal |
|---|---|---|

B2_3

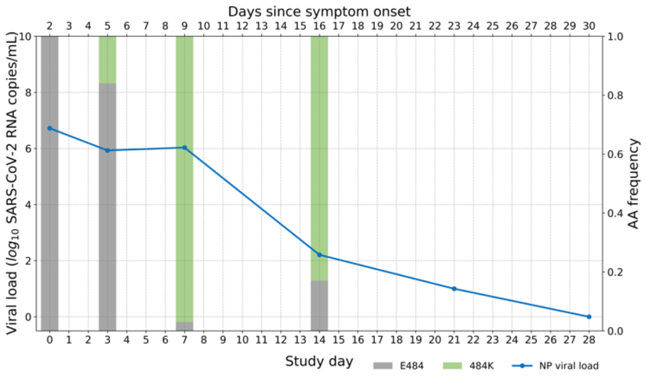


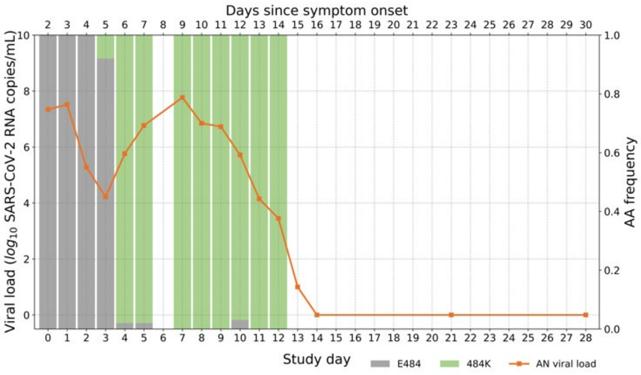


B2_4

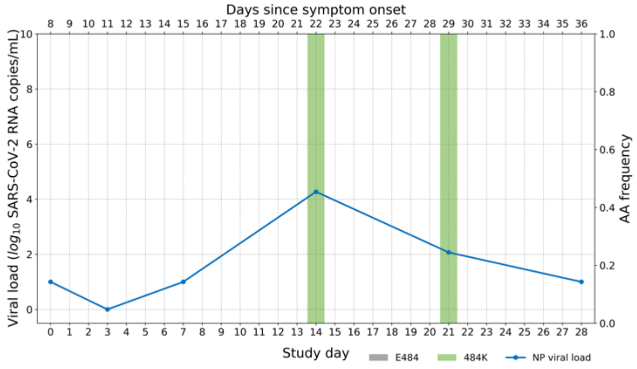


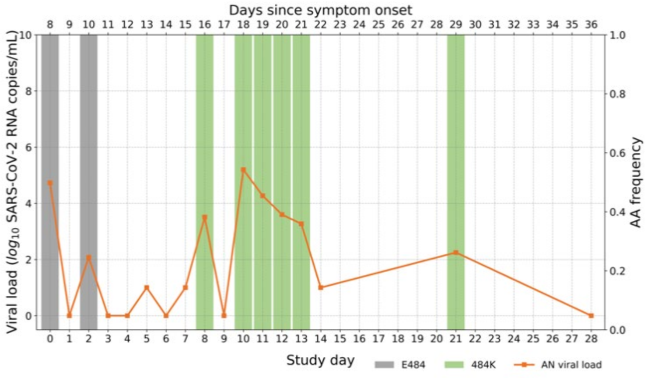

Participant ID | Nasopharyngeal | Anterior Nasal

B2_5

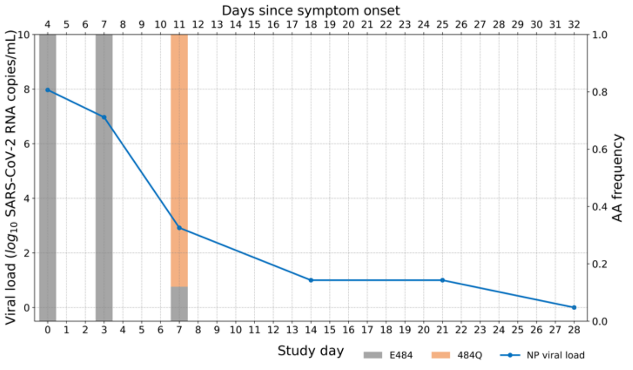


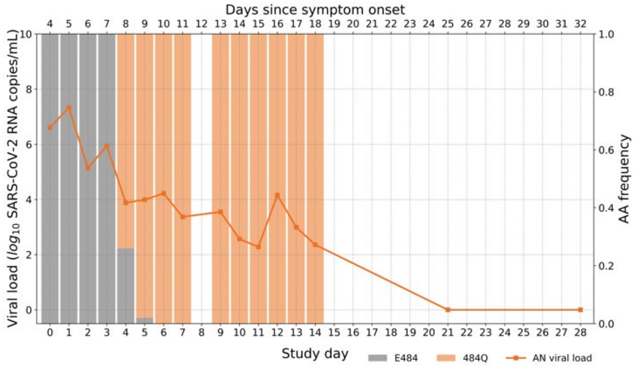


B2_6

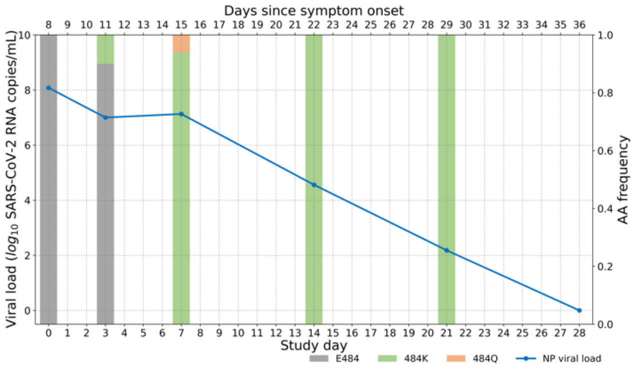


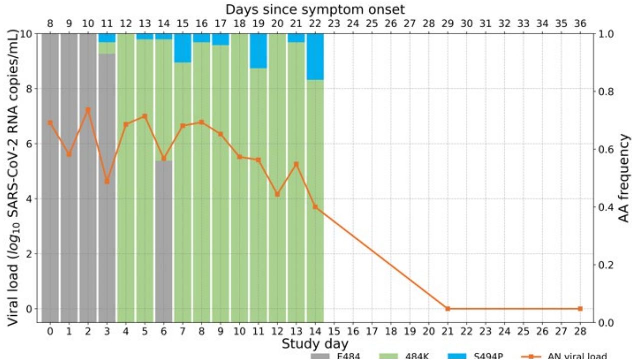


B2_7

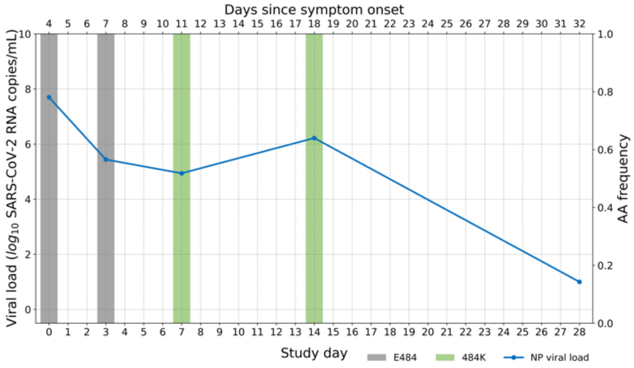


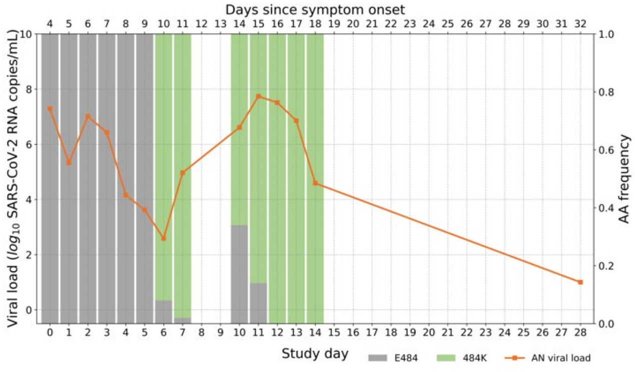


B2_8

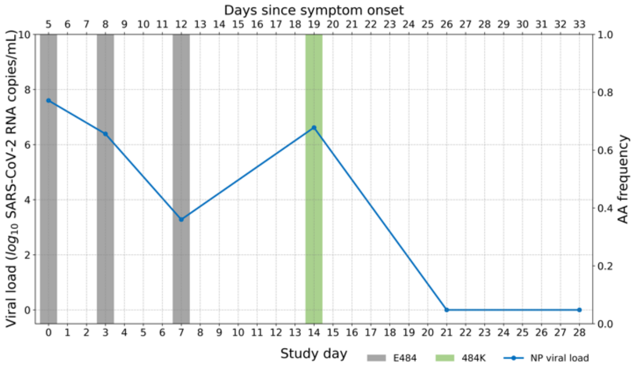


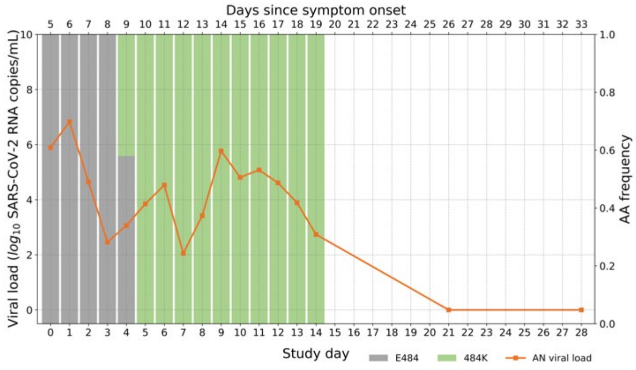

Participant ID | Nasopharyngeal | Anterior Nasal

B2_10

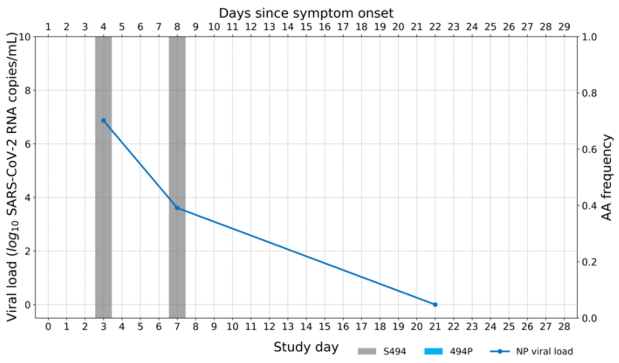


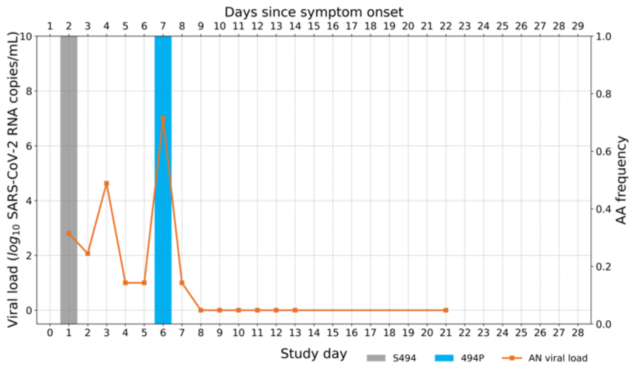


### Treatment arm participant with both baseline and emerging resistance

Participant ID | Nasopharyngeal | Anterior Nasal

B2_2

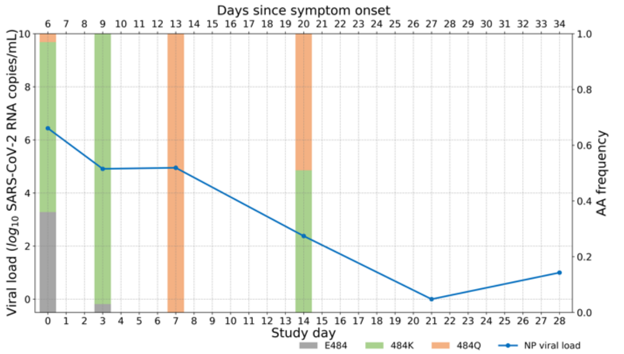


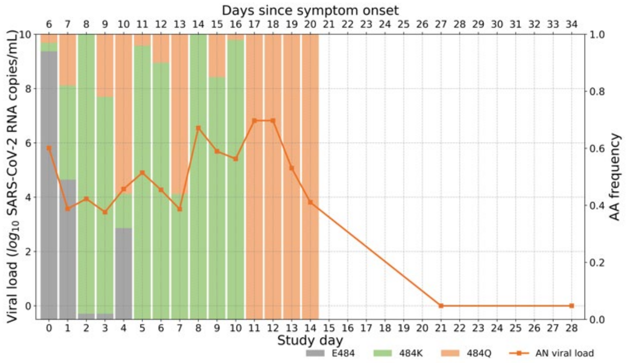

**Supplemental Figure 2:** SARS-CoV-2 specific IgG antibody profiling at baseline in different study groups. Horizontal bars represent median antibody titer. Dashed lines represent antibody positivity detection threshold. RBD denotes Receptor binding domain, NTD N-terminal domain, N Nucleocapsid.

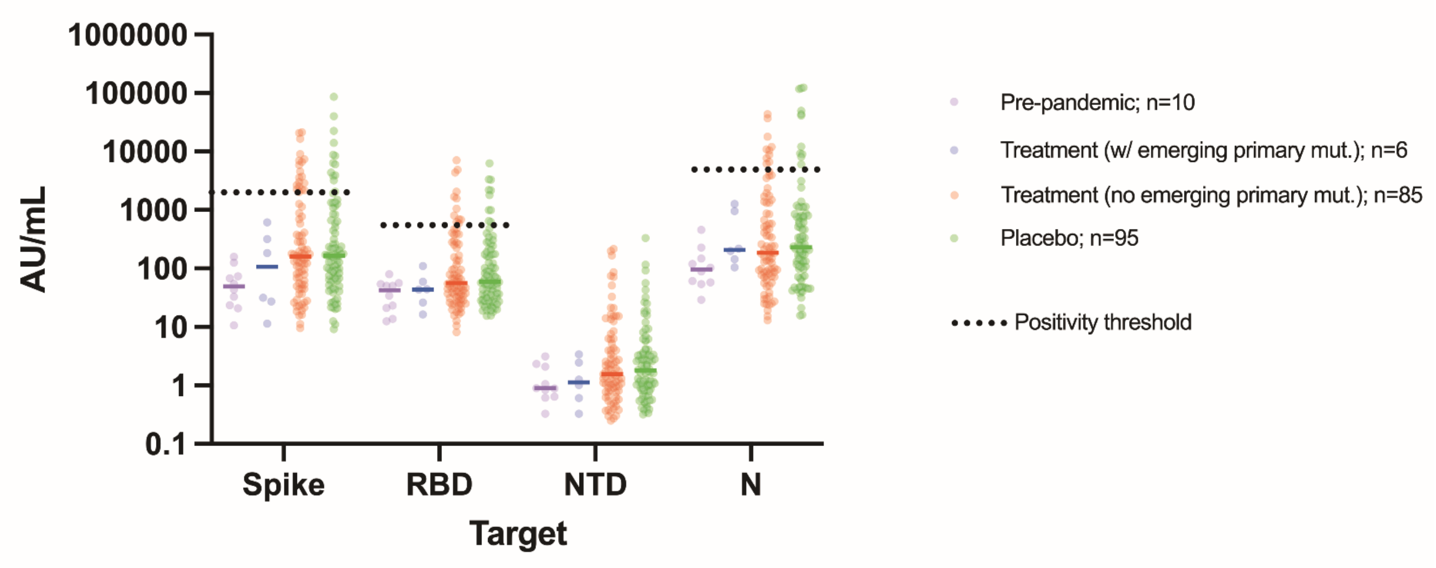

**Supplemental Figure 3: Fitting of the mathematical model to viral load and viral frequency data in individuals with resistance mutations.** (A-F) In each panel, the upper plot shows the viral load kinetics in the individual with the ID shown in the title; the lower plot shows the frequencies over time for the mutants under analysis. Data used for model fitting are shown as 'o' and data not used for model fitting are shown as 'x'. Simulation results using the best-fit parameters (Supplemental Table 2) are shown as lines. (G) Comparison of growth rates of the E484 and the 484K strains estimated from mathematical models for 5 individuals. P-value is calculated using a Wilcoxon signed rank test for paired data.

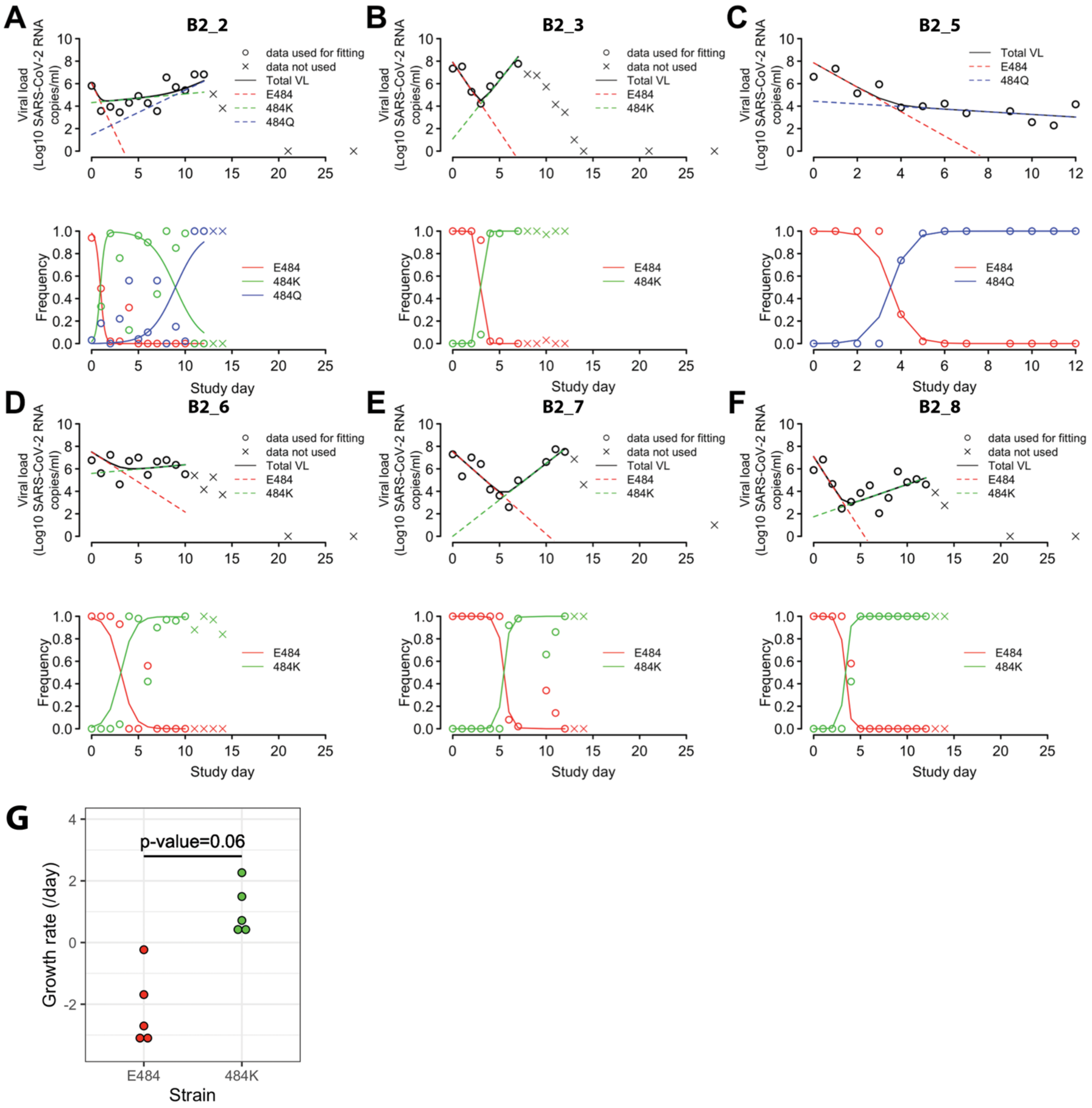

**Supplemental Figure 4: Emerging polymorphisms in different participant counts.** Counts of emerging polymorphisms (including primary resistance sites) in NP samples on day 7 in three study groups: participants with emerging primary resistance mutations, treatment group participants without emerging primary resistance mutations, and the placebo group. Box plots show median and interquartile range.

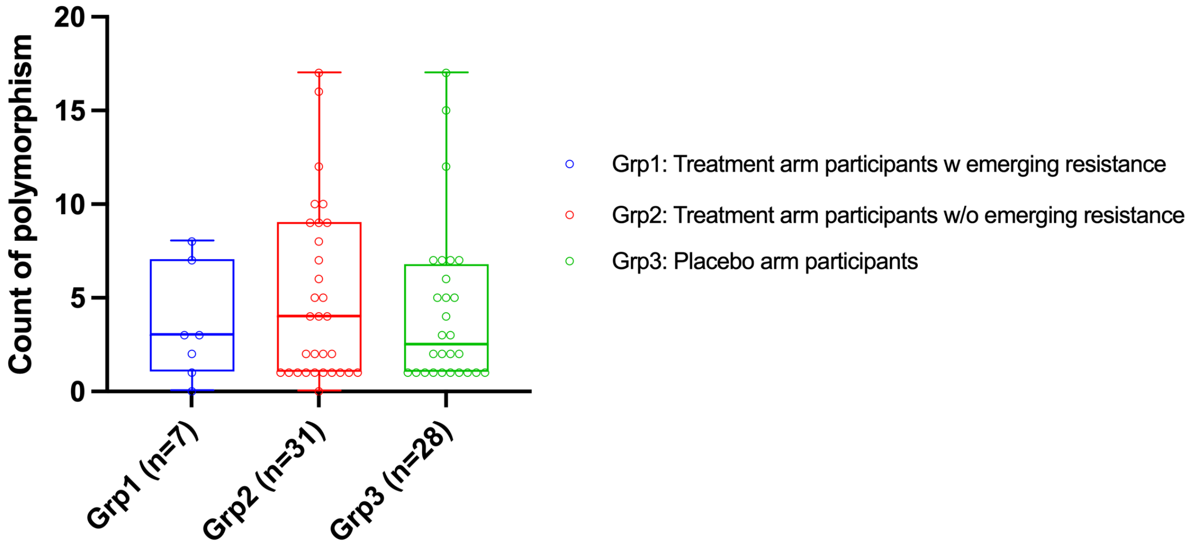

**Supplemental Table 1.** Pharmacokinetic parameters for bamlanivimab in 700mg treatment arm patients with emerging primary resistance mutations. Also included are mean values provided for each parameter by the manufacturer in the U.S. FDA EUA fact sheet for bamlanivimab and etesevimab. Pharmacokinetic data were not available for B2_10. $C_{max}$ was not available for B2_7. $C_{28d}$ for B2_6 was below the limit of quantitation and the value shown was imputed as one-fourth of the lowest quantified concentration. $T_{1/2}$, elimination half-life; $C_{max}$, maximum concentration; $C_{28d}$, concentration at day 28; CL, clearance.

| **Participant ID** | **$T_{1/2}$ (days)** | **$C_{max}$ (μg/mL)** | **$C_{28d}$ (μg/mL)** | **CL (L/day)** |
|---|---|---|---|---|
| B2_2 | 10.00 | 261.13 | 15.74 | 0.31 |
| B2_3 | 19.01 | 243.46 | 35.91 | 0.20 |
| B2_4 | 14.86 | 334.22 | 37.51 | 0.18 |
| B2_5 | 22.24 | 228.21 | 37.02 | 0.20 |
| B2_6 | 4.65 | 164.21 | *2.54 | 0.51 |
| B2_7 | 12.66 | N/A | 37.12 | 0.57 |
| B2_8 | 16.71 | 143.92 | 21.41 | 0.30 |
| **Median** | **14.86** | **235.83** | **35.91** | **0.30** |
| EUA value | 17.6 | 196 | 22 | 0.27 |

**Supplemental Table 2.** The estimated growth rate and frequency at the time of treatment for each mutant based on data from six individuals.

| ID | Growth rate under treatment (per day) | | | Frequency | | |
|---|---|---|---|---|---|---|
| | **E484** | **484K** | **484Q** | **E484** | **484K** | **484Q** |
| B2_2 | -4.1 | 0.2 | 0.9 | 0.98 | 0.01 | 2.5E-05 |
| B2_3 | -2.8 | 2.4 | NA | 1 | 1.6e-07 | NA |
| B2_5 | -2.5 | NA | -0.3 | 1 | NA | 0.0004 |
| B2_6 | -1.2 | 0.2 | 0.6 | 0.99 | 0.01 | NA |
| B2_7 | -1.7 | 1.5 | NA | 1 | 3.0E-08 | NA |
| B2_8 | -3.0 | 0.7 | NA | 1 | 4.5E-06 | NA |
